# Clinical validation of a wireless patch-based polysomnography system: a pilot study

**DOI:** 10.1101/2022.08.04.22278354

**Authors:** F Raschellà, M Knoops-Borm, M Sekeri, D Andries, M Oloo, I Moudab, S Musaka, S Mueller, M Stockhoff, M Tijssen, S Coughlin, H Schneider

**Affiliations:** Onera Health, Eindhoven (Netherlands); Neuro rehab Epilepsy and Parkinson’s Center, Tschugg, (Switzerland); American Sleep Clinic, Frankfurt (Germany); Johns Hopkins University, Baltimore (USA)

## Abstract

**Background:** Polysomnography (PSG) is the gold standard for diagnosing and monitoring sleep disorders, however, it is time-consuming and costly, as the application of the equipment can only be done by trained sleep technicians and the test must be conducted within a sleep laboratory. In this study we assessed the performance of the first wireless patch-based PSG system, the Onera Sleep Test System (STS), which can be applied by the patient and performed outside of the sleep laboratory in settings such as the home. To achieve this, sleep stage and physiological data from the Onera STS were compared to gold standard in-lab PSG.

**Materials and methods:** The recordings were unsupervised to simulate a home-use environment. Epoch-by-epoch agreement was assessed by calculating sensitivity, specificity, accuracy, and Cohen’s kappa coefficient. Pearson’s correlation coefficients were calculated for multiple sleep parameters to measure the level of entire night agreement.

**Results:** Substantial agreement with a Cohens kappa of 0.69 across all sleep stages was determined, which reached 0.81 when Stage N1 was removed from the analysis. A high accuracy, specificity, and sensitivity were found for wake N2, N3 and REM. Although specificity (95.25%) and accuracy (89.62%) were high for N1, sensitivity was low (27.19%). Sleep parameters calculated by sleep stage transitions, apnea hypopnea index and oxygen desaturation index, showed strong correlations.

**Conclusions:** The Onera STS provides comparable clinical information to traditional PSG. Moreover, the application time was reduced by 77% which reduces the overall costs of PSG. These results open the possibility for PSG studies to be performed efficiently outside of the sleep laboratory at a larger scale, thus improving access for patients.

## Introduction

**Polysomnography (PSG) is the gold standard for diagnosing and monitoring sleep disorders. For conducting a PSG, the patient sleeps for a single night in a specialized laboratory, during which a variety of physiological signals, e.g**., **sleep stages and breathing patterns, are measured. This procedure is expensive and time consuming, limiting the number of patients that can get this diagnostic test. The efficiency of sleep diagnostics will increase when self-applied, wireless PSG can be used at home**.

Attended PSG is the gold standard for the diagnosis and management of patients with suspected sleep disorders [1, 2]. During a PSG study, a minimum of 4-hours of quality assured sleep data are collected from patients who sleep in a controlled environment which includes on-line video and vital signs safety monitoring. The high-cost and difficulty recruiting trained, licensed, sleep technicians to conduct this test limits capacity to specialized centers and leaves many patients unable to access it [3, 4]. Ambulatory PSG has been proposed as a viable solution for monitoring sleep outside of the sleep laboratory [5]. However, when using currently available equipment the preparation for the study, including the application of the equipment, still needs to be performed by a trained professional. As a result, unattended PSGs are rarely performed in clinical routine.

Over the last two decades, Home Sleep Testing (HST) has allowed patients with a high probability of severe obstructive sleep apnea (OSA) to be diagnosed, unsupervised, outside of the sleep laboratory [6]. These devices only record a subset of physiological signals, providing less-detailed information to sleep clinicians and often do not allow access to the raw data [7]. However, due to the high false negative and positive rates of this testing method [8]. PSG is still required for obtaining a reliable diagnosis in a large proportion of patients. Additionally, guidelines state that patients with other sleep disorders and those with significant co-morbidities also continue to require PSG [9].

Onera Health has developed the first wireless patch-based PSG system – the Onera Sleep Test System (STS). The Onera STS was designed to overcome the limitations associated with unattended, home, PSG by measuring the full montage of signals required for accurate sleep monitoring and the correct evaluation of physiological changes that occur during sleep. Furthermore, The Onera STS can be used in home (Home healthcare environment) as well as professional Healthcare Facilities. It consists of four body-worn patches, eliminating the need for individual leads and separate electrodes which increases the probability of measuring a patient’s natural sleep. The Onera STS was granted the CE mark in 2021, and FDA clearance in 2022.

The goal of this exploratory study was to compare the performance of the Onera STS to gold standard in-lab PSG, in a controlled environment.

## Materials and methods

### Study Objective

The objective of this study was to compare outcomes between the Onera STS gold standard in-lab PSG, in a controlled environment. We hypothesized that:

1. the Onera STS provides similar sleep and respiratory metrics in comparison to standard PSG
2. the Onera STS significantly reduces the setup time compared to standard PSG.

### Study Design

This was a prospective single center pilot study including 32 subjects referred for a routine sleep study at the American Sleep Clinic, Frankfurt, Germany. The study started on the 21st of January 2022 and was conducted over a four-week period. Subjects who met the inclusion criteria and consented to participate underwent an unsupervised single night sleep evaluation using both the Onera STS system and a standard PSG system. Standard PSG were performed using Embla S4500 devices (Natus Medical Incorporated, USA) and the setup consisted of EEG registration (C3, C4, F3, F4, O1, and O2), left and right EOG (E1-M2 and E2-M1), chin EMG (anterior, left, and right positioning), airflow (oronasal thermistor and nasal pressure), blood oxygen saturation, ECG, tibialis EMG in both legs, breathing efforts (abdomen and thorax), sleeping position, and audio and video recording of the entire night. Recordings were analyzed using RemLogic software (version 1.1, Embla Systems LCC, USA) by trained technicians in accordance with the scoring rules defined by the American Academy of Sleep Medicine version 2.4, 2017 [9].

### Inclusion and Exclusion Criteria

Clinical site personnel reviewed consecutive patient’s medical history for eligibility. Subjects were considered eligible if they were 18 years or older and had been referred to the study center for a routine sleep study. Subjects were excluded if they were unable to provide informed consent, had a history of adhesive skin allergies, had a severe skin condition at the sites of patch administration, had an implanted cardiac stimulator or diaphragmatic pacer, or had an anatomical abnormality that in the opinion of the Principal Investigator made the subject ineligible for inclusion.

### The Onera STS

The Onera STS is a wearable system for measuring physiological signals during a sleep study conducted in the sleep laboratory or at home. The Onera STS performs electroencephalography (EEG), electrooculography (EOG), electromyography (EMG) of the chin and tibialis muscle, and electrocardiography (ECG) and measures respiratory signals, nasal pressure, oxygen saturation (SpO_2_), activity, body position, and snoring loudness via four sensor patches applied to the forehead, upper chest, abdomen, and lower leg as can be seen in Fig. 1. Each sensor consisted of a disposable patch and re-usable pod.

**Fig. 1.**
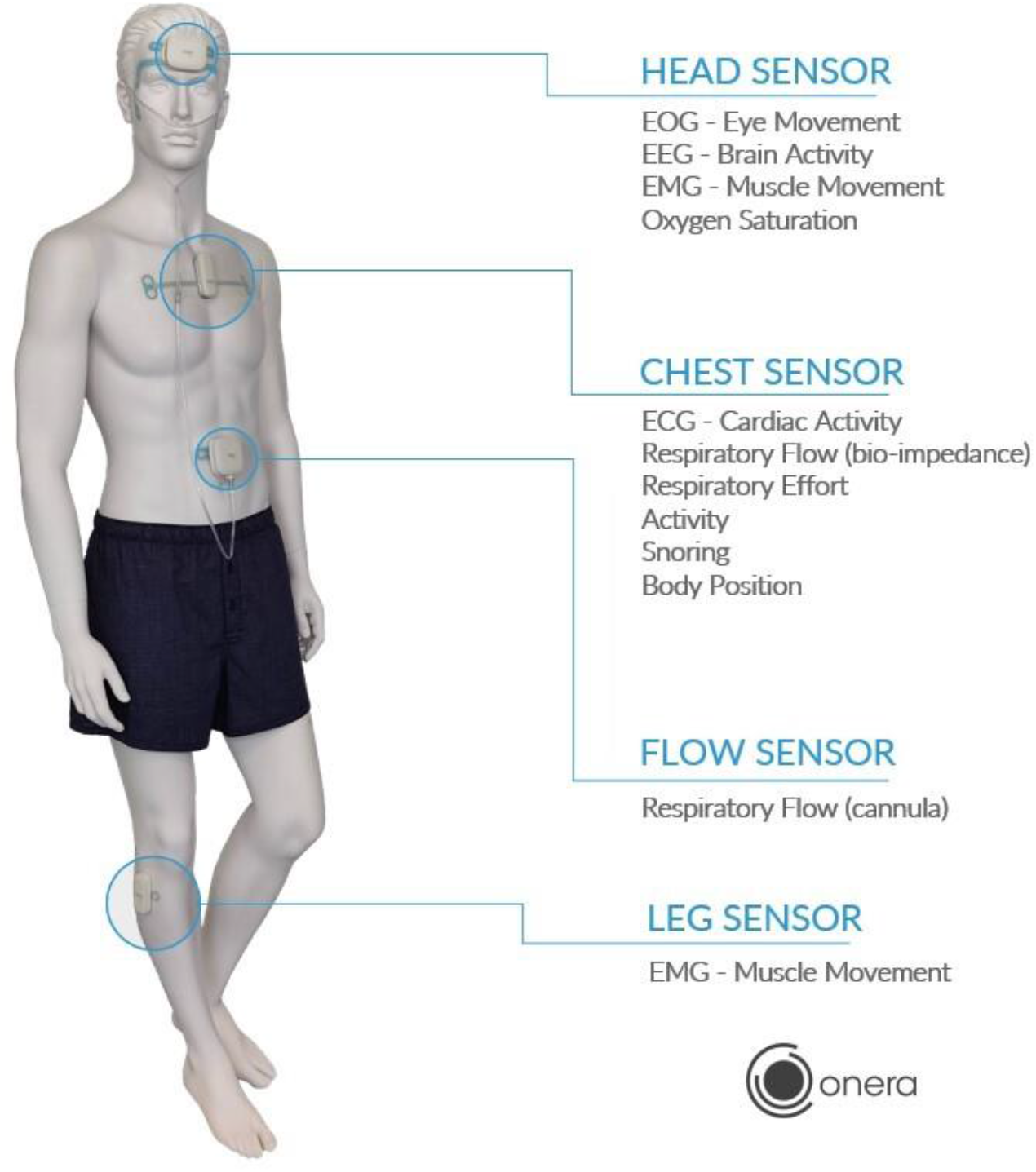
A mannequin wearing the Onera STS, consisting of four sensors: the head, chest, flow, and leg sensor. Each sensor consists of an adhesive hydrogel called ‘the patch’ and a recording unit called ‘the pod’.

### Study Procedures

The Onera STS sensors were applied concurrently to patients by a licensed sleep technician. During application, the set-up time for each device was measured. At bedtime, both devices were activated, and the subject was allowed to go to sleep without further supervision to simulate a home environment. Following completion of the night recording, the data from the Onera STS and PSG were collected. Any studies in which either the Onera STS sensor or the PSG had < 6 hours of recorded data or < 4 hours of data recorded during sleep were excluded from the analysis. Both studies were scored independently using commercially available software by a single qualified sleep professional, according to AASM guidelines [9]. The professional performing the scoring was blinded as to which subjects’ study that they were scoring.

### Clinical interpretation of Onera STS data

The signals recorded by the Onera STS were obtained using a different electrode placement to traditional PSG. Recording examples from data of two Onera study subjects are provided (Fig. 2 and 3) that show the signal morphology and clinical relevance of these signals. These examples show the signals relevant for scoring sleep (Fig. 2) and for detecting respiratory events (Fig. 3). The example of traces of the head patch (Fig. 2) show that a clear distinction can be made between different sleep stages. Alpha activity can be seen in the EEG and EOG signals, and the dissipation of alpha activity, the theta activity together with slow eye movements are visible, both constituting the NREM 1 phase. For classifying NREM stage 2, the data shows typical spindles and K complexes. The data also shows slow wave activity in EEG and EOG signals thus constituting NREM 3. While rapid eye movement can be seen in the EOG signals, they are not visible in the EEG signals which allows the scorer to depict the typical EEG pattern for REM sleep. For the patient represented in Fig. 2, a diagnosis was made of insomnia with hypopneas during transitional sleep stages and REM (rapid eye movement) sleep (normal), when also including the additional respiratory data from the chest patch to calculate the AHI and ODI. In Fig. 3 changes in respiratory flow and effort and the corresponding blood oxygen saturation can be seen, as well as other respiratory parameters. For the patient represented in Fig. 3, the combination of signals provided by the Onera STS allowed the detection of all relevant sleep disordered breathing events. The airflow signal of the bio-impedance flow signal shows several markers of severe upper airway obstruction. The bio-impedance flow signal distinctly shows inspiratory flow limitation in form of 1) an early inspiratory airflow peak, 2) a negative effort dependency (flow is decreasing while effort is increasing) and 3) a prolongation of the inspiratory duty cycle, which are markers previously reported as indicators for inspiratory flow limitation [10]. Addition of the sound signal can differentiate whether periods of upper airway obstruction are associated with no significant sound or with loud snoring. Central or obstructive breathing events as seen in column 3-5 of Fig. 3 can be depicted by the combination of bio-impedance flow and effort and the snoring signal, because of the visible decrease in amplitude of the bio-impedance flow signal.

**Fig. 2.**
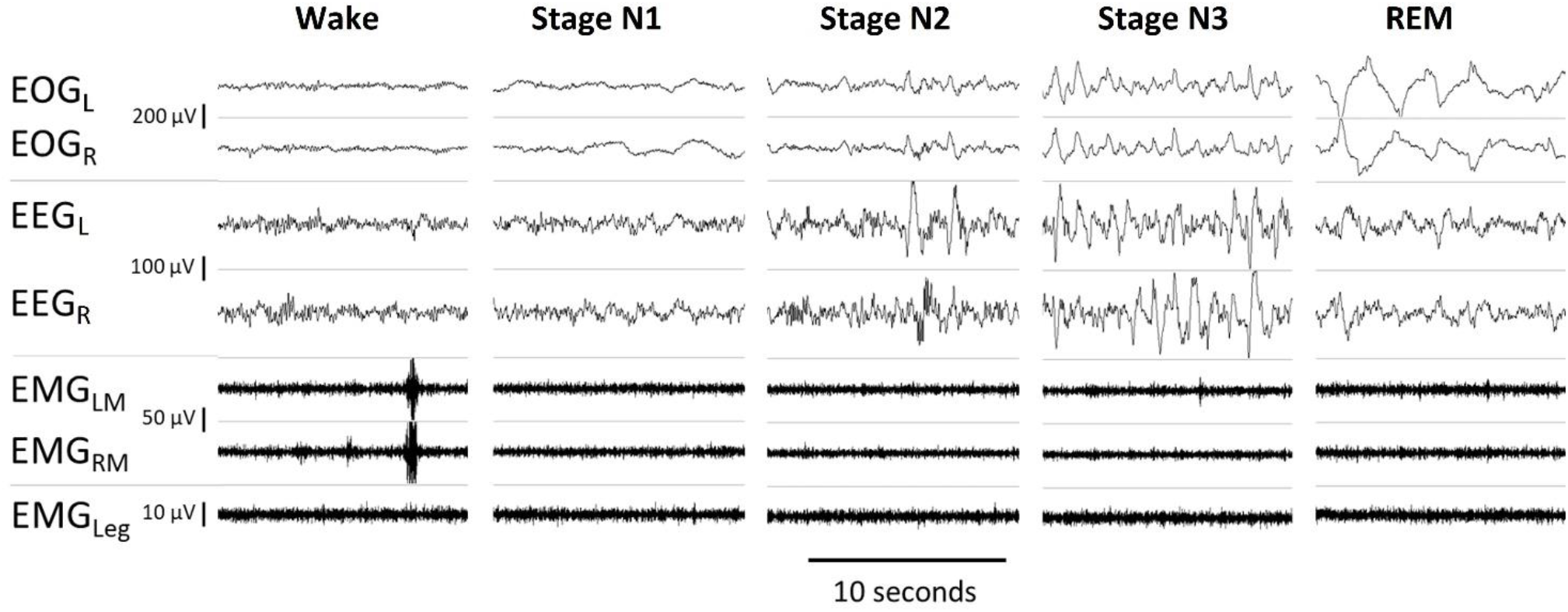
Signals recorded by the Onera STS sensors from a female in her 50s with main complaint of insomnia and a BMI of 21.7 kg/m^2^. 2For all sleep stages, signals show EOG of the left and right eye, EEG of left and right prefrontal lobe, EMG of left and right musculus masseter and EMG of musculus tibialis. The scale of the signals is indicated on the left side.

**Fig. 3.**
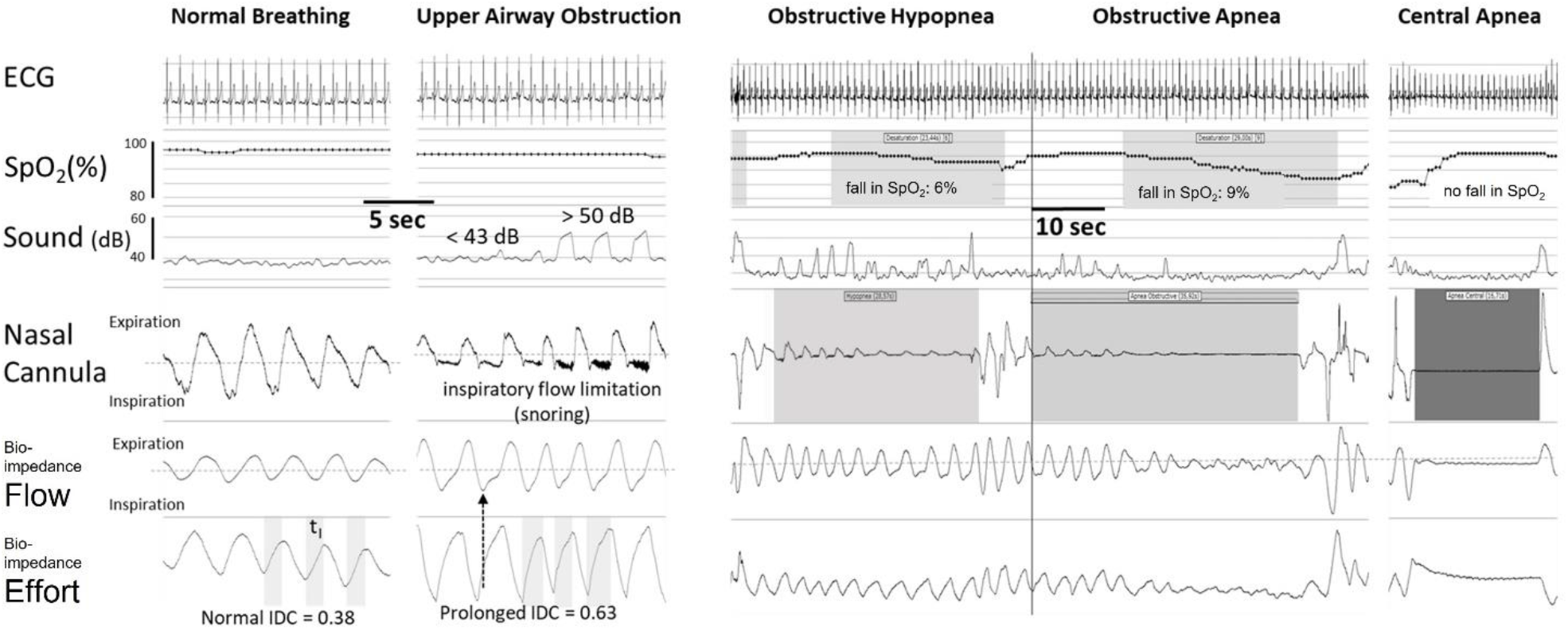
Respiratory events measured from the Onera STS showing the breathing signals of a male in his 40s with main complaint of daytime hypersomnia and a BMI 33.8 kg/m^2^. From left to right, episodes of normal breathing, upper airway obstruction, obstructive hypopnea, obstructive apnea, and central apnea breathing events with corresponding ECG, SpO_2_, sound, respiratory flow (bio-impedance flow) and respiratory effort (bio-impedance effort) signals and nasal flow from the nasal cannula can be seen. In the SpO_2_ signal, desaturations are indicated with corresponding apnea, obstructive apnea, and central apnea in the signal from the nasal cannula. In the second panel from right please note that the amplitude of the bio-impedance flow signal is less than 30% of surrounding breaths, thus constituting an apnea, compared to the nasal cannula signal and bio-impedance flow signal on the far-right panel.

### Outcomes

The clock time was used to synchronize the sleep staging of the Onera STS and PSG for comparative analysis. The following parameters were calculated; Total Sleep Time, wake time, time in N1, N2, N3, REM and NREM, Sleep Efficiency (SE), Sleep Onset Latency (SOL), Wake After Sleep Onset (WASO), REM Onset Latency (ROL), Apnea Hypopnea Index (AHI), SPO_2_ and Oxygen Desaturation Index (ODI).

### Statistical analysis

Epoch-by-epoch agreement for all sleep stage between the Onera STS and the PSG was assessed by calculating accuracy, sensitivity, specificity, and Cohen’s kappa. The Cohen’s kappa coefficient was calculated to determine the level of entire night agreement. A kappa value of 0–0.2 was considered slight agreement, 0.21–0.4 fair agreement, 0.41–0.6 moderate agreement, 0.61–0.8 substantial agreement and >0.8 almost perfect agreement [11].

Pearson’s correlation coefficients were calculated over the duration of all sleep stages to evaluate the agreement between the two devices. As correlation coefficients may be misleading about the level of agreement when comparing clinical measures, Bland–Altman plots are shown for all sleep stages [12]. Finally, the similarity in all sleep parameters was evaluated between the two devices using a paired t-test. Multiple comparisons were adjusted by applying the sequential rejective Bonferroni procedure of Holm [13].

All statistical analyses were performed using MATLAB (9.6.0.1472908 (R2019a) Update 9, The MathWorks Inc., Natick, Massachusetts, USA).

## Results

### Study population

In total, 32 patients were consented into the study. They were middle-aged, predominantly male, overweight and had a range of comorbidities. The patient characteristics can be found in Tab. 1.

**Tab. 1.** Patient characteristics

| Characteristic |  |
| --- | --- |
| Age (years) | 44.9±2.5<sup>a</sup> |
| Male, n (%) | 29 (90.6) |
| BMI (kg/m<sup>2</sup>) | 27.2±0.8<sup>a</sup> |
| Hypertension, n (%) | 5 (15.6) |
| Cardiac Arrhythmia, n (%) | 4 (12.5) |
| Pulmonary Diagnosis, n (%) | 3 (9.4) |

<sup>a</sup> Mean ± standard deviation (unless stated otherwise)

Fourteen studies had <6 hours of recorded data or <4 hours of data recorded during sleep and were excluded from the analysis. From these, the proportion that failed for technical reasons were similar between the devices (approximately 35 % on each device) and in line with technical failure rates reported with other marketed HST devices [14]. The remaining 18 studies were used for analysis.

### Application time

Set-up time for both the Onera STS and the standard PSG were measured by the technician in 28 of the 32 patients (87.5 %) (Tab. 2). Average set-up time for the PSG was 27 minutes and 28 seconds (minimum 14 minutes, maximum 50 minutes), compared to 6 minutes 23 seconds (minimum 3 minutes 37 seconds, maximum 16 minutes, and 42 seconds) for the Onera STS. On average, application time was reduced by 21 minutes and 5 seconds when using the Onera STS, an absolute reduction of 77%.

**Tab. 2.** Application time in minutes for Onera STS and predicate device

|  | Onera STS | PSG |
| --- | --- | --- |
| Mean | 6:23 | 27:28 |
| Standard deviation | 3:12 | 9:19 |
| Min | 3:37 | 14:00 |
| Max | 16:42 | 50:00 |

### Sleep stage performance

Overall, the Onera STS demonstrates high accuracy across all stages (table 3). Sensitivity and specificity were high for wake, N2, N3 and REM. Although specificity was >95% for Stage N1, sensitivity was low. A substantial overall scoring agreement between Onera STS, and PSG based on an overall kappa score of 0.69. When Stage N1 was removed from the analysis, this kappa increased to 0.81.

**Tab. 3.** Epoch-by-epoch comparison between Onera STS and predicate device. Accuracy, sensitivity, and specificity are shown as mean ± standard deviation.

|  | Accuracy (%) | Specificity (%) | Sensitivity (%) |
| --- | --- | --- | --- |
| Wake | 94.08 ± 4.34 | 97.87 ± 2.00 | 61.92 ± 21.70 |
| NREM Stage 1 | 89.62 ± 4.23 | 95.25 ± 3.02 | 27.19 ± 12.11 |
| NREM Stage 2 | 84.69 ± 4.29 | 81.55 ± 8.05 | 88.32 ± 4.80 |
| NREM Stage 3 | 95.60 ± 1.80 | 98.26 ± 2.05 | 76.60 ± 18.58 |
| REM | 94.70 ± 3.27 | 95.95 ± 2.66 | 88.12 ± 14.46 |

Further comparison between the Onera STS and PSG was made by calculating Pearson’s correlation coefficient and performing Bland-Altman analysis across the duration of all sleep stages. This comparison can be seen in Fig. 4. The correlations found range between 0.64 and 0.91 which shows a fair to good correlation between the two devices. The lowest correlation was found for wake and the highest for REM. The Bland-Altman plots show the difference in duration of each sleep stage between the two devices (y-axis) and the mean duration over all measurement (x-axis). The upper and lower limits of agreements are indicated by dotted lines. The interval widths of all sleep stages are similar and range between 35 to 65 minutes, with the smallest interval for N1 and the highest interval for N3 and wake. The bias values (dark line) are around zero for all sleep stages indicating similar performance for both devices. A larger spread of sample for N2 was found due to the low sample size.

**Fig. 4.**
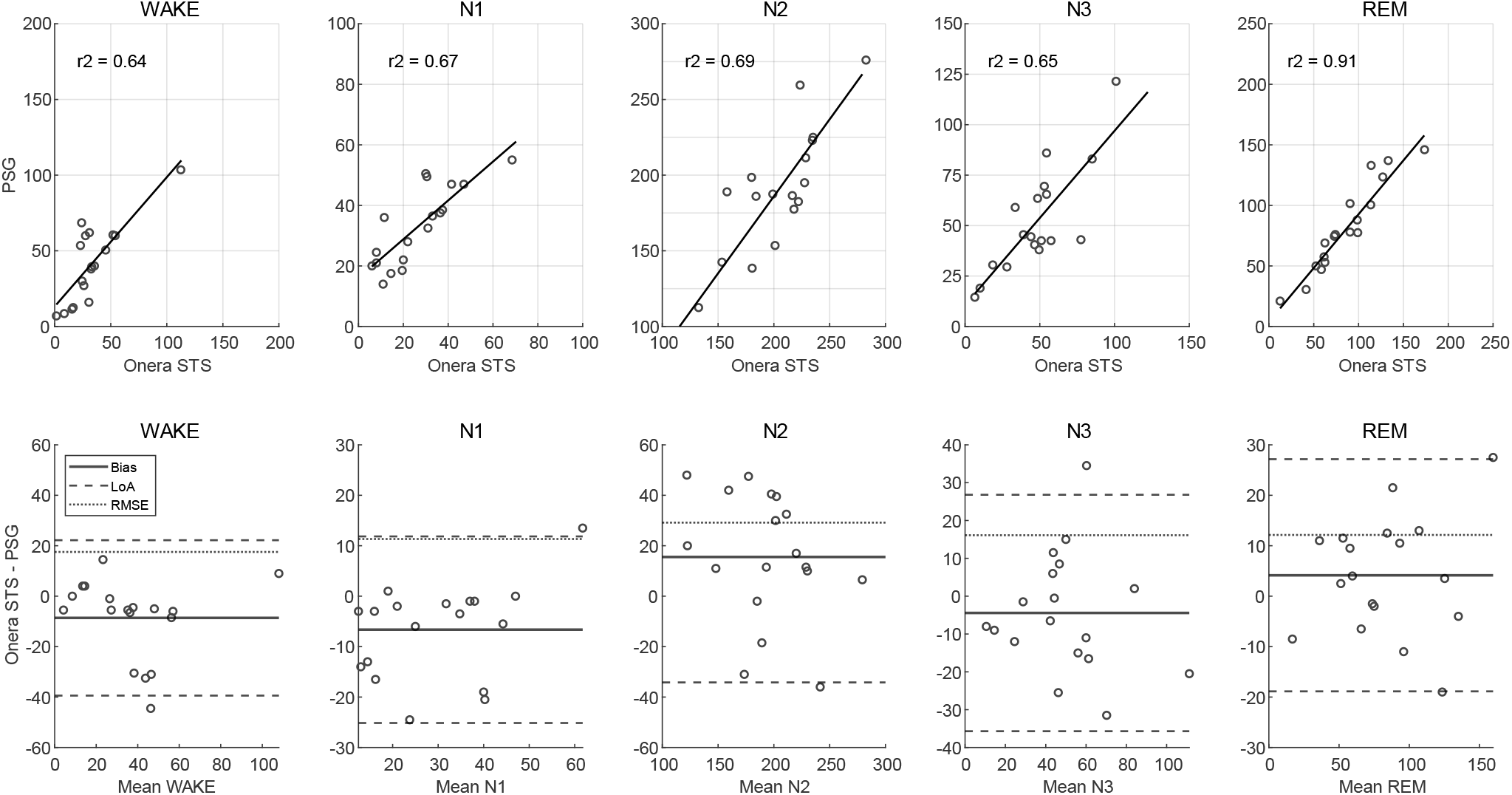
Comparison of duration of sleep stages (in minutes) between the Onera STS and the PSG. In the top panels scatterplots with linear regression line (solid line) and in bottom panels Bland–Altman plots are shown. For the Bland–Altman plots, difference in duration was determined by subtracting the duration of each sleep stage determined by Onera STS from the PSG. Average duration is the mean over all measurements of the Onera STS and PSG. The bias value is shown with the solid line, the limit of agreements (LoA) is shown with dashed lines, and the root mean square error (RMSE) is shown with dotted lines.

### Sleep variables performance

Sleep study outcome measures were compared between the two devices by performing a t-test (table 4). These data shows that the scored variables from Onera STS were not statically different from the PSG for most of the parameters. A significant difference between the devices was found for the Total Sleep Time, although the individual sleep stages showed no difference and the SpO_2_. However, this difference was so small that it did not lead to a statistically significant difference in the apnea hypopnea index or oxygen desaturation index.

**Tab. 4.** Average sleep parameters over all patients for traditional PSG versus Onera STS in minutes (as mean ± standard error of the mean) unless stated otherwise and corresponding p-value.

| Parameter | PSG | Onera STS | p |
| --- | --- | --- | --- |
| TST<sup>a</sup> | 374.4±10.8 | 396.7±9.7 | 0.0006 |
| Wake | 62.1±9.3 | 73.2±9.4 | NS<sup>g</sup> |
| NREM Stage 1 | 38.2±3.3 | 33.7±4.0 | NS |
| NREM Stage 2 | 197.1±9.1 | 216.9±7.0 | NS |
| NREM Stage 3 | 53.2±6.1 | 49.2±5.7 | NS |
| REM<sup>b</sup> | 86.2±8.9 | 96.9±8.7 | NS |
| NREM | 288.5±10.1 | 299.8±8.7 | NS |
| REM (%) | 22.8±2.1 | 24.2±1.9 | NS |
| NREM (%) | 77.2±2.1 | 75.8±1.9 | NS |
| SE (%)<sup>c</sup> | 86.0±1.9 | 83.8±1.9 | NS |
| SOL<sup>d</sup> | 45.6±6.4 | 45.9±6.2 | NS |
| WASO<sup>e</sup> | 39.4±6.4 | 27.3±5.8 | NS |
| ROL<sup>f</sup> | 105.4±10.4 | 103.6±8.9 | NS |
| AHI (events/hour) | 19.7±4.2 | 19.7±3.8 | NS |
| SpO<sub>2</sub> (%) | 94.1±0.3 | 95.4±0.3 | 0.0019 |
| ODI (events/hour) | 13.6±3.8 | 15.2±3.1 | NS |

<sup>a</sup> Total Sleep Time, <sup>b</sup> Rapid Eye Movement, <sup>c</sup> Sleep Efficiency, <sup>d</sup> Sleep Onset Latency, <sup>e</sup> Wake After Sleep Onset, <sup>f</sup> REM Onset Latency, <sup>g</sup> Not Significant

## Discussion

In this study, we have demonstrated that the Onera STS had a similar performance on sleep and respiratory parameters when compared to gold standard PSG and that the application time of the Onera STS is 77% lower than that of a PSG. The concordance between the scored sleep and respiratory parameters from the Onera STS and PSG suggest that the data traces recorded by the Onera STS had signal characteristics and resolution that was adequate for analysis by an experienced sleep scorer.

We noted minor differences in the agreement in NREM 1 between the devices, which we believe arose either due to the low sample size of individuals or the lower time spent in NREM 1 compared to other sleep stages. Regardless, a lower intra-rater agreement for scoring NREM 1 is common and thus comparable to previous head-to-head comparisons of PSG systems. We do not believe this was due to differences in the placement of the EEG electrode, because we have observed that the Onera STS EEG signals clearly depict alpha activity (Fig. 2) and WASO and REM sleep, which both rely on the recognition of alpha activity, were similar. Thus, we believe that once a larger sample size for validation is acquired, we will observe increased agreement for NREM1.

An intriguing, but unexpected, finding of this study was the statistically small difference found in concurrently recorded SpO_2_ values, which were higher when obtained at the forehead by the Onera STS than obtained from the finger by the PSG. We believe that this may be due to either, changes in the peripheral vasculature of the patients or technical differences between the SpO_2_ sensors such as the sampling rate and artifact removal algorithms. Although not reported here, the PI of this study was able to replicate this finding using a Nonin (Nonin Medical Inc., USA) head and finger sensor suggesting that physiologically, the lower SpO_2_ at the finger could be related to REM specific vasoconstriction of small arterioles of the fingertip or to a fall in arterial blood pressure (nocturnal dipping) below a threshold that triggers counter-active vasoconstriction in the periphery to preserve circulation of central organs. If correct, this hypothesis could be important for disease recognition.

The data in this study were scored by one, independent, accredited, scorer. As all studies were scored by this individual, we eliminated inter-scorer variability and the bias of inexperience. Conversely, there were two main limitations. Firstly, a small sample size, predominance of males and limited age-range means these data are not generalizable to a normal sleep laboratory population. The small sample of patients may also have led to a misrepresentation of comorbidities that could influence physiological signal integrity during a sleep study. Therefore, we suggest that future studies include a larger number of subjects so that gender, age, and comorbidities are more accurately represented. Secondly, as studies were not supervised by a technician telemetrically, there were failed in laboratory studies, which is not usual for an in-laboratory workflow, and which further reduced the sample size for analysis.

A robust patch-based PSG system which can be applied by patients and perform studies in the sleep laboratory and home environment may have significant positive clinical and economic impacts on the healthcare system and improve the patients experience of a sleep study. Currently, a PSG relies on a sleep technician to attach electrodes and leads to the patients to transfer physiological signals to the hardware for recording and analysis, and an overnight stay in the sleep laboratory. A patch-based PSG system that can be applied more quickly by the sleep technician or can be self-applied by patients would significantly decrease the engagement of the clinical team whose time can be focused on other tasks and mean that sleep labs are no longer confined by the limited availability of skilled technicians. Similarly, performing the test at the patients’ home would significantly reduce the number of sleep technicians required for overnight supervision and cost for an overnight stay in the sleep laboratory. Moreover, the ability to perform the test at home allows patients to sleep more naturally and provide a more representative overview of their sleep. Another, benefit of the Onera STS, compared to almost all HST devices, is the ability to measure other sleep disorders besides sleep disordered breathing outside of the sleep laboratory environment. Thereby, the Onera STS expands clinicians’ ability by to detect other sleep disorders, such as the disruption in sleep architecture,. which is beneficial for patients with psychiatric and neurological conditions and may have a significant impact on both the evolution and management of these conditions.

## Conclusion

In this study, we have demonstrated that the Onera STS had a similar performance on sleep and respiratory parameters when compared to traditional PSG and that the application time of the Onera STS is 77% lower than that of a PSG which will lower the burden on the sleep technicians and reduce the overall operational costs of PSG. Therefore, Onera STS will open the possibility for PSG studies to be performed more efficiently in a sleep center and potentially opens the opportunity to extend it outside of the clinical setting at a larger scale, thus improving access for a broader proportion of patients.

### Practical conclusion

- We developed a self-applied, wireless, PSG system which can be used at home
- The Onera STS device performs similarly to traditional PSG and will improve the efficiency of sleep diagnostics in the future
- The improved application time of the Onera STS will lower the burden on the sleep technician

## Data Availability

All data produced in the present work are contained in the manuscript.

## Compliance with ethics guidelines

### Conflict of interest

M. Stockhoff, M. Knoops-Borm and M. Tijssen are current employees of Onera Health. M. Knoops-Borm has shareholding options in the company. Until recently, F. Raschellà and M. Sekeri were employees of Onera Health. H. Schneider and S. Coughlin are contractors for Onera Health, receive compensation and have shareholdings in the company. H. Schneider receives compensation for studies conducted at the American Sleep Clinic. All other authors report no conflicts of interest.

The study was managed by an independent CRO (Maxis Medical Inc, Frankfurt), who ensured regulatory compliance and compliance with ethical board requirements to prevent bias from COI.

### Patients’ rights protection statement

All procedures followed were in accordance with the ethical standards of the committee responsible on human experimentation (institutional and national) and with the Helsinki Declaration of 1975 (in its most recently amended version). Informed consent was obtained from all patients included in the study.

### Ethics

The clinical trial described in this manuscript was registered with an internationally recognized trial registry (ClinicalTrials.gov ID: NCT05310708).

